# FORESEE: A prospective feasibility trial of functional precision oncology for metastatic breast cancer patients

**DOI:** 10.64898/2026.10.10.26365226

**Authors:** Christos Vaklavas, Saundra S. Buys, Sandra D. Scherer, Kristen Kelley, Ling Zhao, Chieh-Hsiang Yang, Emilio Cortes-Sanchez, Andrew Butterfield, Xiaomeng Huang, Philip J. Moos, Yi Qiao, Jake Reed, Kenneth M. Boucher, Gabor Marth, Bryan E. Welm, Alana L. Welm

## Abstract

The FORESEE trial (NCT04450706) was a prospective study to evaluate the feasibility of integrating organoid-based drug testing with comprehensive genomic characterization to inform treatment selection in patients with HER2-negative metastatic breast cancer. The primary objective of the study was to determine the feasibility of providing personalized therapy recommendations using a combination of genomic vulnerability information with drug testing in patient-derived organoids (PDOs) in a clinically relevant time frame. Of the 15 patients enrolled, 10 cases had biopsies with adequate tumor content, and PDOs were successfully established in 4 of 10 cases. Drug profiling and genomic testing was completed in all established organoids, yielding an overall success rate of 40% in cases of biopsies with adequate tumor content. Drug sensitivity and genomic results were returned within 12 weeks and informed subsequent treatment selection in all evaluable cases. The primary endpoint - demonstrating that PDO-based drug screening combined with genomic characterization would identify clinically actionable outcomes in at least 46% of patients based on prior data - was not met. In conclusion, successful PDO establishment represents a limiting step in the implementation of FPO for metastatic breast cancer. These findings highlight the need for improved strategies optimizing use of patient biopsy material for drug testing to fully realize the clinical potential of FPO.

## Introduction

Metastatic breast cancer (MBC) remains incurable, and with an estimated 42,140 deaths projected in the United States in 2026 (American Cancer Society, Siegel et al., 2026) there is a pressing need to develop more effective therapeutic strategies. More than 40 unique drugs have been approved by the FDA for the treatment of breast cancer. Although these therapies have demonstrated efficacy at the population level, breast cancer is a highly heterogeneous disease, and only a limited subset of treatments is likely to provide meaningful clinical benefit to any individual patient. There are adverse effects associated with systemic therapies, and each line of therapy can increase treatment resistance and narrow eligibility options for subsequent therapies. Thus, accurately matching the most effective therapy to each patient’s tumor as early in the course of treatment as possible, while minimizing exposure to ineffective treatments, is a critical clinical priority and the central goal of precision oncology.

The current precision oncology paradigm is centered on identification of genomic driver alterations by next-generation sequencing (NGS) and choosing drugs that target the respective gene product or pathway. This concept constituted a paradigm shift in cancer therapeutics (Gladstone et al., 2025, Schwartzberg et al., 2017). However, most cancers do not harbor actionable genomic alterations, and even when such alterations are identified, it remains uncertain whether target inhibition will translate into clinically meaningful benefit (André et al., 2014, reviewed in Letai et al., 2022). In the NCI-MATCH trial, 37.6% of patients were found to harbor actionable alterations, but only 1 7.8% were assigned to a matched therapy protocol, even with all 38 treatment arms activated. The overall response rate among evaluable patients was only 1 0.3%, albeit with significant variability of activity among and within treatment arms (Flaherty et al., 2023; O’Dwyer et al., 2023). Tumors, especially metastases, evolve in complex microenvironments, and their intrinsic heterogeneity and plasticity enables dynamic adaptation in response to therapeutic pressures. These properties represent major challenges for effective treatment selection and are not adequately captured by static genomic snapshots (Letai et al., 2022). Furthermore, genomic profiling typically informs only the selection of particular classes of targeted therapies. Chemotherapy remains the therapeutic mainstay for most patients with metastatic cancer, especially for later lines of therapy. In metastatic breast cancer, the use of antibodies with cytotoxic payloads (antibody drug conjugates) is rapidly expanding and enables targeted delivery of chemotherapy to kill tumor cells, but only works if the tumor cells are sensitive to the payload.

Patient-derived models have emerged as important tools in translational cancer research, drug discovery and development (Pauli et al., 2017)). Unlike conventional cell line models, patient-derived xenografts (PDXs) and organoids (PDOs) retain most of the molecular, genomic, and phenotypic characteristics of the originating tumor cells, allowing for more accurate predictions of therapy response - and they can be used on a personalized level for functional precision oncology (FPO). The central tenet of FPO is the incorporation of a functional test(s), often alongside static measurements like genomic mutation testing, to more accurately predict tumor drug responses and personalize therapy (Letai et al., 2022). Indeed, studies have shown remarkable one-to-one concordance in terms of drug activity between the models and originating tumor (Hidalgo et al., 2014, Zhang et al., 2013). PDXs and PDOs can be established from limited input material, such as tumor biopsies (Sachs et al., 2018, Verstegen et al., 2025) and PDO drug profiling studies can be completed within clinically applicable time frames that are not always feasible with PDX models. We and others have shown that organoids retain fidelity to the originating tumor (Sachs et al., 201 8; Chen et al., 2021) and drug responses in organoids have been validated in matching PDXs (Guillen et al., 2022). In a sentinel case of metastatic triple negative breast cancer, we showed that organoid-based drug screening studies can meaningfully inform treatment selection (Guillen et al., 2022). PDOs have been utilized in clinically oriented studies in breast (Wu et al., 2024; Zheng et al., 2025), colorectal (Ooft et al., 2021; Jensen et al., 2023), and prostate cancer (reviewed in Pamarthy and Sabaawy 2021), underscoring their potential for integration into clinical practice workflows.

Our FPO platform extends the standard precision oncology paradigm beyond static genomic assessment by integrating PDO-based drug response profiling with comprehensive tumor genomic data comprising single cell or single nucleus RNA-sequencing (RNA-seq) and whole genome sequencing (WGS). We designed the FORESEE trial (Functional Precision Oncology for Metastatic Breast Cancer; NCT04450706) to prospectively and systematically evaluate the clinical utility of this FPO platform. The primary objective was to investigate the feasibility of integrating such testing into clinical practice to inform treatment selection in metastatic breast cancer. Secondary objectives included assessing the turnaround time for returning results to treating physicians and evaluating the performance and concordance of the integrated functional precision oncology (FPO) approach relative to commercial genomic testing platforms. Exploratory objectives were to assess the efficacy of FPO-informed treatments compared to the prior line of therapy, and to compare which treatment nominations arose with FPO versus with standard commercial circulating tumor (ct)DNA and/or tissue genomic testing.

### Study design

Eligible patients were women with newly diagnosed metastatic triple negative (TNBC) or hormone receptor (estrogen receptor and/or progesterone receptor) positive/HER2 negative (HR+/HER2-) breast cancer who had exhausted and/or expected to derive no further clinical benefit from endocrine therapy options (Figure 1, upper panel). Patients with TNBC whose disease relapsed early (<12 months after completion of treatments with curative intent) were eligible and prior adjuvant or neoadjuvant therapy did not constitute a prior therapy for the purposes of the trial. Additional key inclusion criteria were willingness and capability to undergo tumor biopsy from a metastatic site, and histologic confirmation of the biopsy as ⍰ 20% tumor content. Patients needed to have an estimated life expectancy of ⍰ 6 months. After the enrollment of the first 3 patients (part 1), enrollment was paused to allow for preliminary review of feasibility and implementation of changes to optimize workflow. Part 2 of the trial enrolled 12 additional patients, equally divided between TNBC and HR+/HER2- (Figure 1, upper panel).

**Figure 1.**
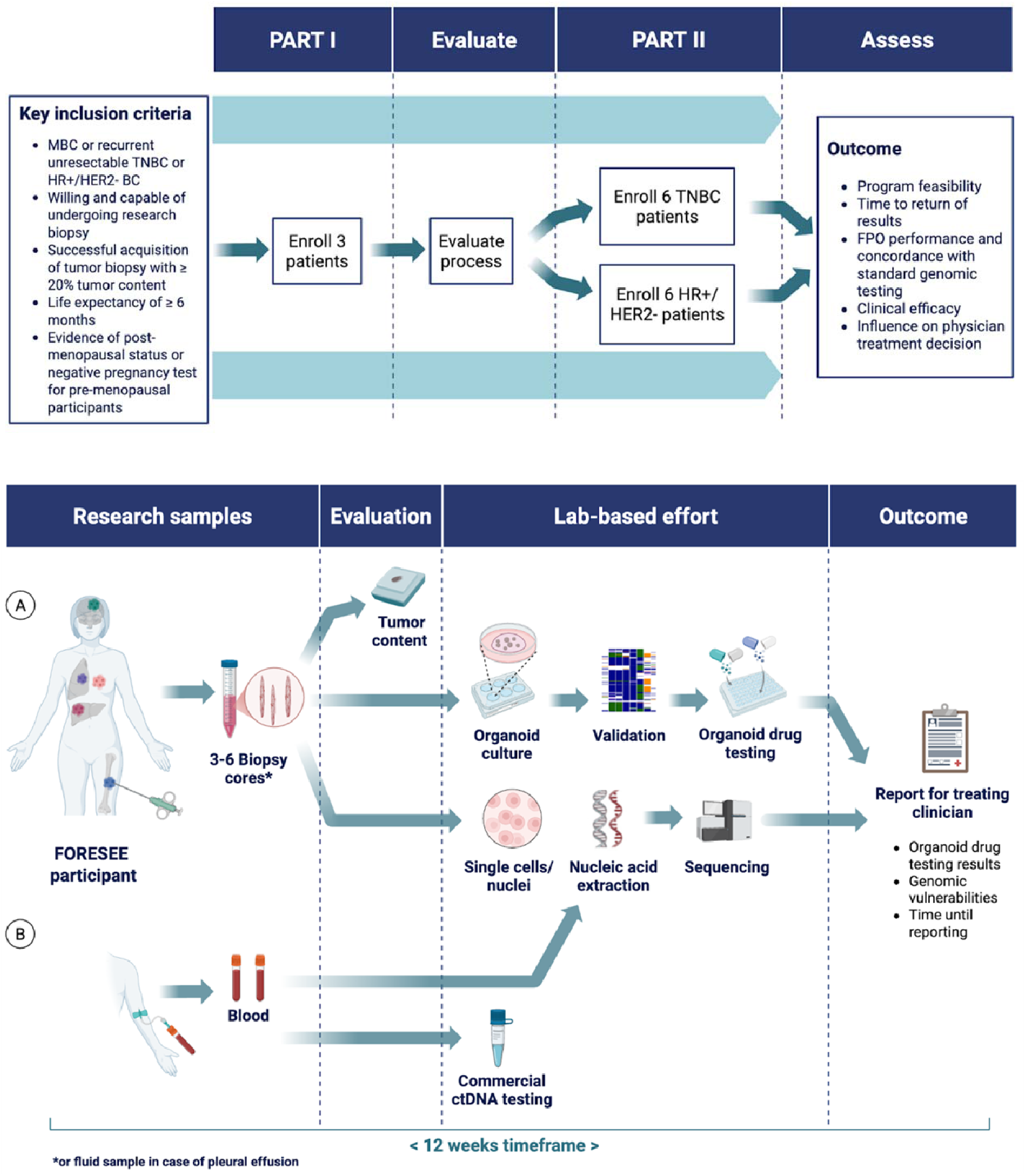
Study Design (upper panel) and Study Procedures (lower panel). Tumor tissue or fluid sample was acquired from eligible patients for PDO establishment and genomics (A), and blood for germline DNA and ctDNA (B). After the biopsy was performed, patients began therapy at the discretion of the treating physician. The genomics results from ctDNA and tumor tissue were compiled along with the results from PDO drug screening in a single report that was shared and discussed with the treating physician within 12 weeks. **Created in** https://BioRender.com

### Study Procedures

Study procedures entailed a research biopsy of a metastatic site and blood collections (Figure 1, lower panel). Biopsy tissue was evaluated for tumor content by a board-certified pathologist and used for single cell or single nuclei RNA sequencing (scRNA-seq/snRNA-seq), WGS, and PDO development for drug screening. scRNA-seq/snRNA-seq and WGS constituted collectively the *investigational tumor sequencing*. Patients went on to receive their first/next line of systemic therapy at the discretion of the treating physician, and progression-free survival (PFS1) time was recorded. In TNBC and HR+/HER2-metastatic breast cancer, the first line therapeutic options have been well articulated in phase 3 clinical trials (Cortes et al., 2022; Bardia et al., 2024). Meanwhile, tumor organoids were cultured to establish PDO lines. Validated PDOs (see below) were subjected to dose-response testing with drugs that are FDA-approved for breast cancer, as well as investigational agents available to the patient in active ongoing clinical trials within the institution. Even though a very broad range of drugs and combinations thereof can be evaluated in our FPO platform, the emphasis of this trial was on candidate therapies that could be readily available to the patient, to evaluate the immediate clinical utility of the platform.

The results from the investigational tumor sequencing were compiled along with results from PDO drug screening in a single report that was shared and reviewed with the treating physician. The subsequent line of therapy was chosen by the treating physician and could align with the findings in the report (“informed therapy”) or not (“uninformed therapy”); physicians were not required to utilize or follow the FPO data. Response assessments were conducted per standard of care until documented radiographic or clinical progression; the study did not dictate disease assessments at prespecified intervals. Progression-free survival on the 2nd line of therapy (after the return of FPO results, whether it was an “informed therapy” or not) was recorded as PFS2, and the efficacy endpoint was determined as PFS2/PFS1 >1. For the primary endpoint comprising the proportion of cases where clinically meaningful therapeutic vulnerabilities would be identified by our FPO platform within the clinically relevant timeframe of 12 weeks, a one-sided exact binomial test with null hypothesis that the true proportion is 46% or lower was performed.

Key amendments to the protocol upon completion of Part 1 were exclusion of patients whose only site amenable to tumor biopsy was hepatic metastases (though patients with liver metastases were not excluded), due to consistently unsuccessful PDO generation from liver metastases; and inclusion of patients with fluid collections (pleural, pericardial, ascitic) with confirmed presence of tumor cells as source material for organoid development.

## Results - primary objective

In total, 17 patients were assessed for eligibility. Two cases were deemed ineligible; 15 patients were enrolled in FORESEE (TNBC, n = 7; HR+/HER2-, n = 8; recurrent, n = 11; *de novo* metastatic, n = 4) (Figure 2A, Supplemental Table 1). The median age was 55.5 years (HR+/HER2-, 52; TNBC, 57); the median number of prior therapies in the metastatic setting for patients with HR+/HER2-breast cancer was 3.5 with a median duration on therapy immediately prior to enrollment of 2.27 months.

**Figure 2.**
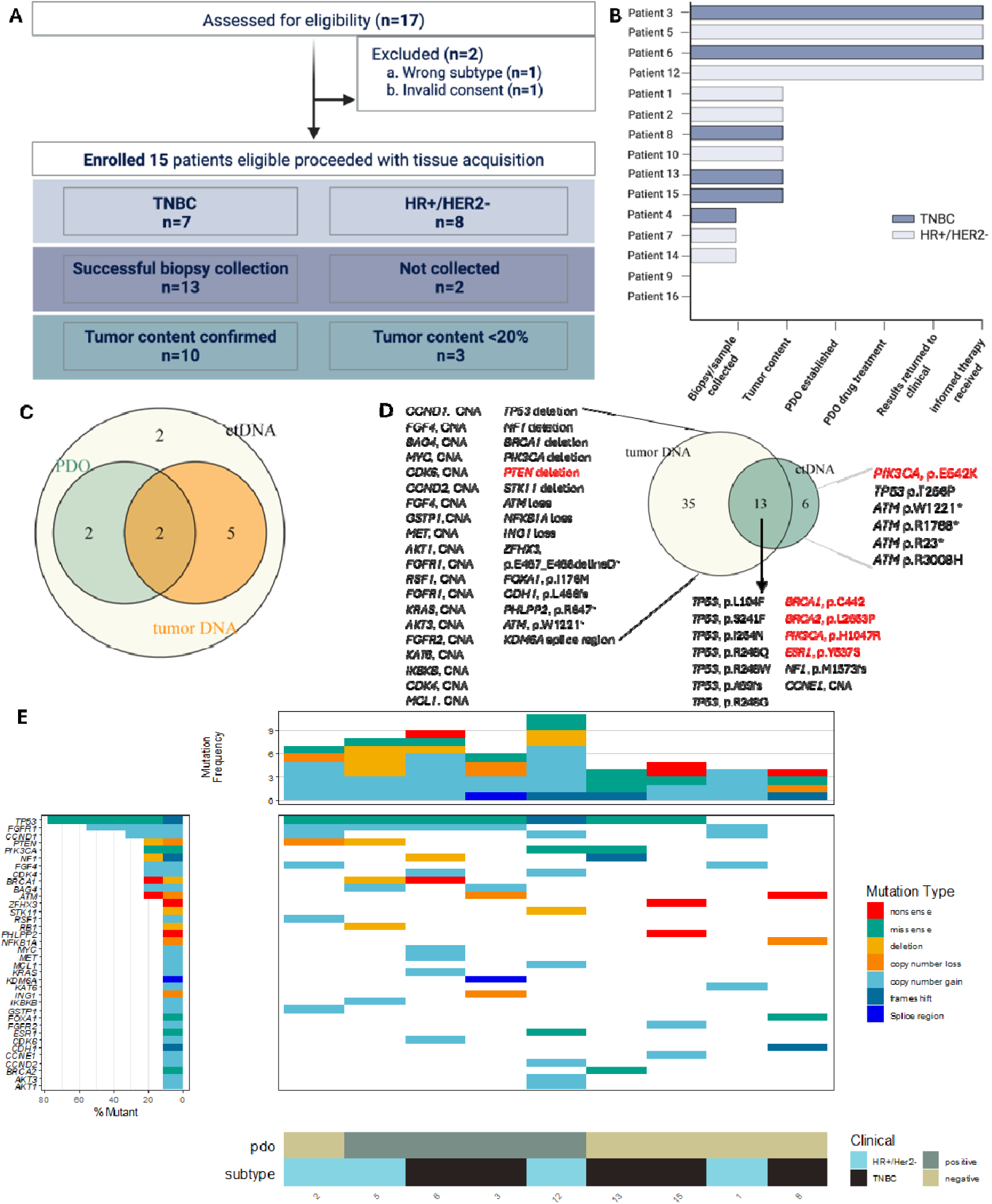
**A**. CONSORT diagram of the FORESEE trial. **B.** Swimmer plot of progress through the FORESEE trial workflow for each enrolled patient. **C**. Venn diagram of the patients who underwent commercial genomic tumor and circulating tumor (ct)DNA testing. All 4 patients in whom PDOs were established underwent investigational tumor tissue DNA testing. **D.** Genomic aberrations detected by ctDNA (commercial) vs. tumor (merged commercial and investigational) DNA testing in the 9 patients who underwent concurrent circulating tumor and tumor tissue DNA testing. Actionable aberrations are highlighted in red. CNA, copy number aberration. E. Genomic aberrations detected in the 9 patients who underwent tumor tissue DNA testing. Cumulative frequencies of genomic aberrations by gene shown on the left, cumulative genomic aberrations per patient shown on top, breast cancer subtype and PDO establishment shown at the bottom.

Biopsies were successfully collected from 13 of 15 patients (87%), with adequate tumor content (>20%) obtained from 10 of 15 patients (67%). PDOs were successfully established and validated for tumor content from 4 of these 10 patients (TNBC, n = 2; HR+/HER2-, n = 2; Figure 2B). PDO drug screening was successfully completed for all 4 validated PDO models and FPO results with candidate treatment nominations were returned within the prespecified 12-week timeframe, yielding a 40% overall success rate (95% CI 12%-74%).

### Events

Pain related to the investigational biopsy (Grade 1, n = 6; Grade 2, n = 3) constituted the most frequent adverse event in the study. Grade 1 post-procedural hemorrhage (n = 1) constituted the second most frequent adverse event related to the trial.

### Candidate treatment nomination by genomics testing

Eleven patients underwent commercial ctDNA testing, 7 of whom underwent concordant commercial tumor DNA testing (Figure 2C). All 4 patients for whom PDOs were established underwent commercial ctDNA testing and investigational tumor DNA sequencing. Note that commercial testing was triggered once patients underwent successful tumor tissue biopsy and tumor tissue was prioritized for PDO generation. For the 9 patients that underwent concurrent ctDNA and tumor DNA testing (commercial, n = 7; investigational, n = 3 [1 patient underwent both]), even though tumor DNA testing revealed many more genomic aberrations than ctDNA testing (most notably copy number aberrations and deletions), most mutations with therapeutic implications were detected by both ctDNA and tumor DNA testing (Figure 2D). The notable exception was *PTEN* deletion, which was detected only on tumor DNA testing (n = 2). In one case, an actionable mutation in *PIK3CA* at low variant allele frequency was detected in ctDNA testing but not in tumor testing; notably, the PDOs generated from this patient were found to be sensitive to the PI3K targeting agent alpelisib (patient 5, discussed below).

The mutational landscape of the 9 patients who underwent tumor DNA testing is shown in Figure 2E. Loss of function mutations in *TP53* (7/9), *FGFR1* copy number gain (5/9), and *cyclin* D1 (*CCND1*) copy number gain (3/9) constituted the most frequent genomic aberrations. Commercial tumor and/or ctDNA testing identified potentially actionable alterations (i.e. protein-altering variants with an associated therapy based on evidence from the medical literature) and nominated targeted treatment options in 5/11 cases (TNBC, 3/6 cases; HR+/HER2-, 2/5 cases). In 2 cases, genomic testing detected and made recommendations for olaparib on the basis of previously known germline mutations in *BRCA2* (n = 1) and *ATM* (n = 1); however, literature does not support the use of olaparib in patients with HER2-breast cancer and *ATM* mutations (Tung et al., 2026). In one case harboring both *ESR1* and *PIK3CA* mutations, elacestrant and fulvestrant with alpelisib were nominated without prioritizing one option over the other (patient 12, discussed below). Lastly, in one case, ctDNA analysis detected an ESR1 mutation, but no treatment was nominated, as none of the novel selective estrogen receptor degraders were approved at that time.

### Candidate treatment nomination by FPO

A treatment was nominated in all 4 cases with successful PDO establishment (hereafter referred to as PDO(+)). Drug response evaluations included not only assessing the absolute activity of each drug on individual PDOs, but also comparing the activity of any given drug across all prior patient-derived models tested, to account for potential differences in assay performance among drugs. We weighed the relative activity of each drug against each tumor, along with the availability of each drug for each patient, considering their tumor mutation status, historic treatment and other clinical factors assessed by the physician on an individualized basis. Thus, the most potent drug was not always the one nominated.

Figure 3 illustrates the natural history of the PDO(+) patients and when across the natural history of their disease the FPO-informed therapy was given. All four PDO(+) participants had recurrent disease. Intervals between completion of treatments with curative intent and histologic diagnosis of recurrence ranged from 0.42 to 4.39 years. Patients with HR+/HER2- breast cancer had relapsed while on adjuvant endocrine therapy prior to trial enrollment. Pertinent PDO-based drug dose-response profiling results that informed treatment selection are shown in Figures 4-7 and are summarized in Table 1.

**Figure 3.**
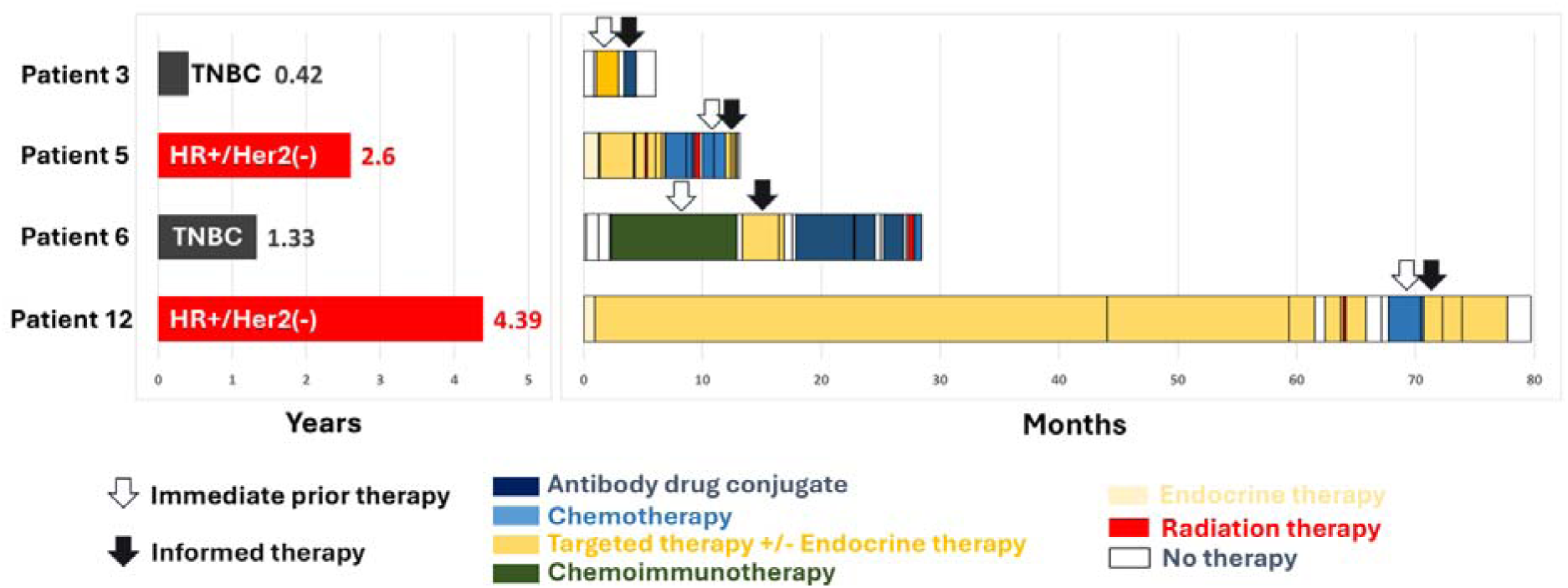
Natural history of the PDO(+) patients and recurrence-free interval**. Left panel**. All PDO(+) patients had recurrent disease and the intervals of remission (interval between completion of all treatment modalities with curative intent (marked as time zero) and diagnosis of recurrent metastatic disease) in years are illustrated on the left bar graph. **Right panel**. Prior treatments received and progression-free intervals in the metastatic setting are illustrated on the right. Time zero constitutes the time of histologic confirmation of metastatic disease. Arrows mark the FPO-informed (solid arrow) and immediate prior (empty arrow) line of therapy.

**Figure 4.**
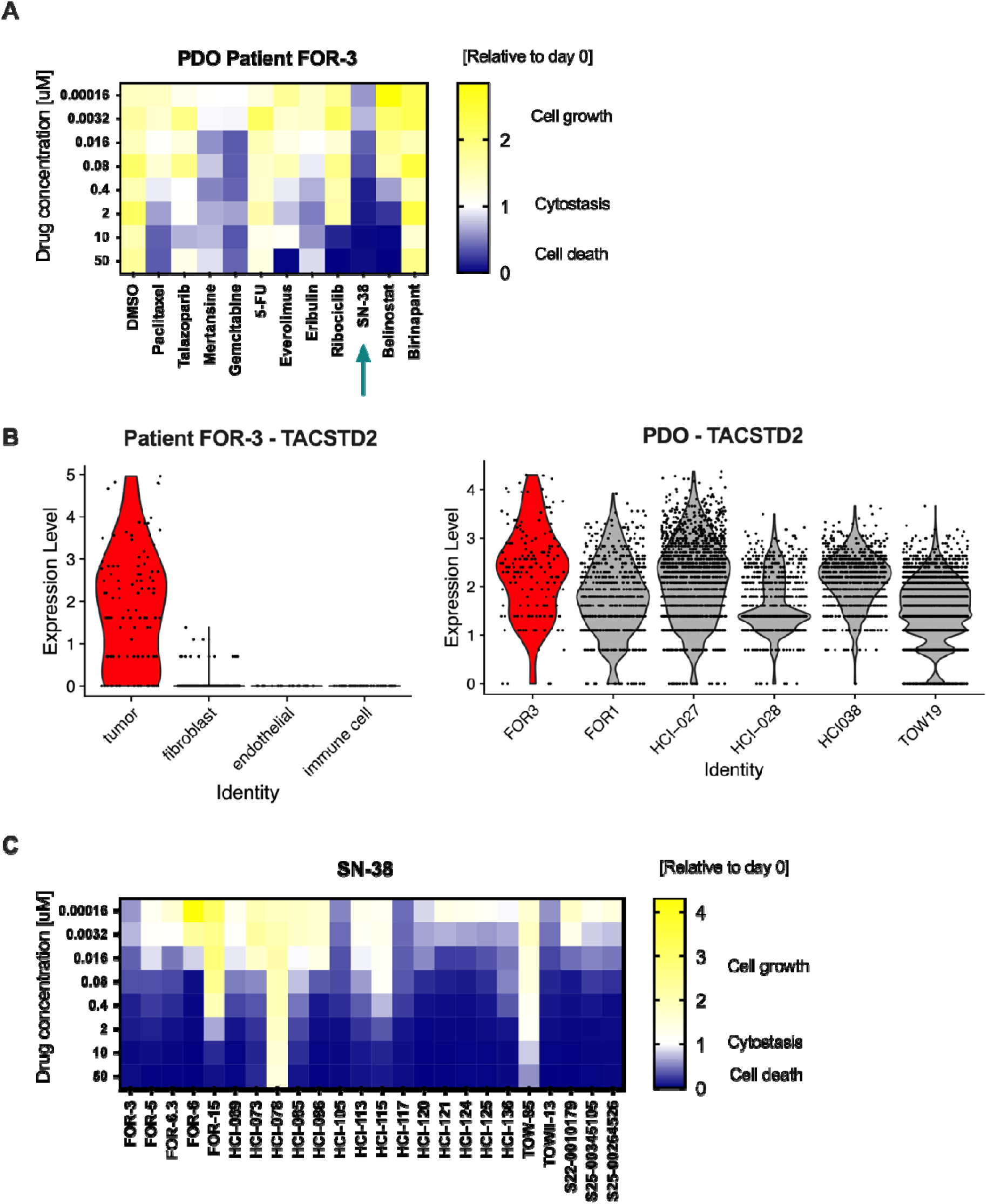
Integrated FPO results that informed treatment selection for patient 3 (FOR-3). (A) PDO drug response assay results for patient FOR-3, showing Cell Titer Glo (CTG)-based ATP levels as a surrogate for viability for each treatment at the indicated concentrations (average of 3 biological replicates, with 4 technical replicates each). For all CTG heatmaps, darker blue indicates cytotoxicity, white indicates a cytostatic response, and yellow indicates growth on drug. The arrow indicates the informed therapy that was nominated. (B) Single cell RNAseq data analysis for *TACSTD2* gene expression as a surrogate for TROP2 in Patient 3 tumor cells versus other cells (left), and compared to tumor cells in other patient-derived models (right). (C) PDO drug sensitivity to SN-38 compared to responses in other patient-derived organoids tested previously. The arrow indicates PDO from patient 3.

**Table 1.** Summary of results, treatment nominations, and outcomes on PDO(+) patients.

| PDO (+)<br>Patient | Actionable<br>target (treatment<br>nomination by<br>commercial<br>genomics) | Treatment<br>nomination<br>by FPO | Clinically relevant findings<br>from investigational genomics | Patient's next<br>treatment &<br>physician's<br>choice | PFS2:PFS1<br>ratio |
| --- | --- | --- | --- | --- | --- |
| **Patient 3** | None (no<br>nomination<br>made) | Sacituzumab<br>govitecan* | Epithelial-mesenchymal<br>transition gene expression<br>signature | Sacituzumab-<br>govitecan | 0.52 |
| **Patient 5** | *PIK3CA* p.E542K<br>(alpelisib) | Alpelisib | Luminal B signature<br>*RB1* loss | Alpelisib &<br>exemestane | 0.26 |
| **Patient 6** | *BRCA1* p.C422*<br>(PARP inhibitor) | Abemaciclib | High copy number gains in<br>*CDK4* and *CDK6* | Abemaciclib &<br>exemestane | 0.29 |
| **Patient 12** | *ESR1* p.Y537S,<br>*PIK3CA*<br>p.H1047R<br>(elacestrant OR<br>fulvestrant +<br>alpelisib) | Everolimus | Additional mutations<br>downstream of the *PIK3CA*<br>predicting lack of response to<br>alpelisib: *AKT1* & *AKT3*<br>amplification; *STK11* loss | Everolimus &<br>exemestane | 0.6 |

\*Nomination made on the basis of sensitivity of the PDO to SN38 and RNA expression levels of the gene encoding TROP2.

Physicians were required to fill out surveys before and after the disclosure of the FPO results. Before disclosure of the results, physicians indicated that having concordant genomic and FPO results would be most influential in terms of their treatment selections. In case of discrepancy between commercial genomic and FPO results, most physicians indicated that the FPO results would carry greater weight. Accordingly, in all cases the results of FPO testing informed the subsequent line of therapy.

In the first PDO(+) case, representing early recurrent metastatic TNBC (Patient 3; Figure 4), neither tumor nor ctDNA sequencing nominated a treatment. Investigational tumor RNA-seq was consistent with a strong epithelial-mesenchymal-transition signature. PDO-based drug screening revealed a significant response to SN-38, and a medium response to gemcitabine (Figure 4A). All other drugs tested showed weak antitumor activity, reflective of the chemotherapy-refractory nature of the patient’s disease. SN-38, the active metabolite of irinotecan, is the cytotoxic payload of the FDA approved anti-TROP2 antibody-drug conjugate sacituzumab govitecan. We interrogated the patient’s tumor gene expression data for the gene encoding TROP2 (*TCSTD2*) and found robust expression levels (Figure 4B). The collective results nominated sacituzumab govitecan as the optimal 2nd line therapy, which also constituted the standard of care at that time. The patient received only one cycle of the antibody drug conjugate before it was withheld due to declining performance status without unequivocal evidence of progressive disease.

In the second PDO(+) case (Patient 5, Figure 5), the treatment (alpelisib) nominated by FPO and ctDNA (by virtue of detection of a *PIK3CA* activating mutation) aligned; tumor DNA testing did not detect a *PIK3CA* mutation (or other actionable mutations) and hence, did not nominate a treatment. Investigational tumor RNA-seq revealed a luminal B-like signature with high expression of *ESR1* and *FOXA1*, and a high proliferation signature. Loss of *RB1* predicted resistance to CDK4/6 inhibitors and correlated with the very short prior progression-free interval with exemestane and palbociclib followed by exemestane and abemaciclib (3 and 2 months, respectively). Even though the PDO’s response to alpelisib was cytostatic rather than cytotoxic, it ranked in the top 1 5% of previous patient-derived models previously tested with this drug (n=47; Figure 5B). There was an average to poor response, relative to other models, to commonly used chemotherapies in this setting (paclitaxel, eribulin), the conjugate SN-38, and other targeted therapies (talazoparib, everolimus, abemaciclib, tucatinib).

**Figure 5.**
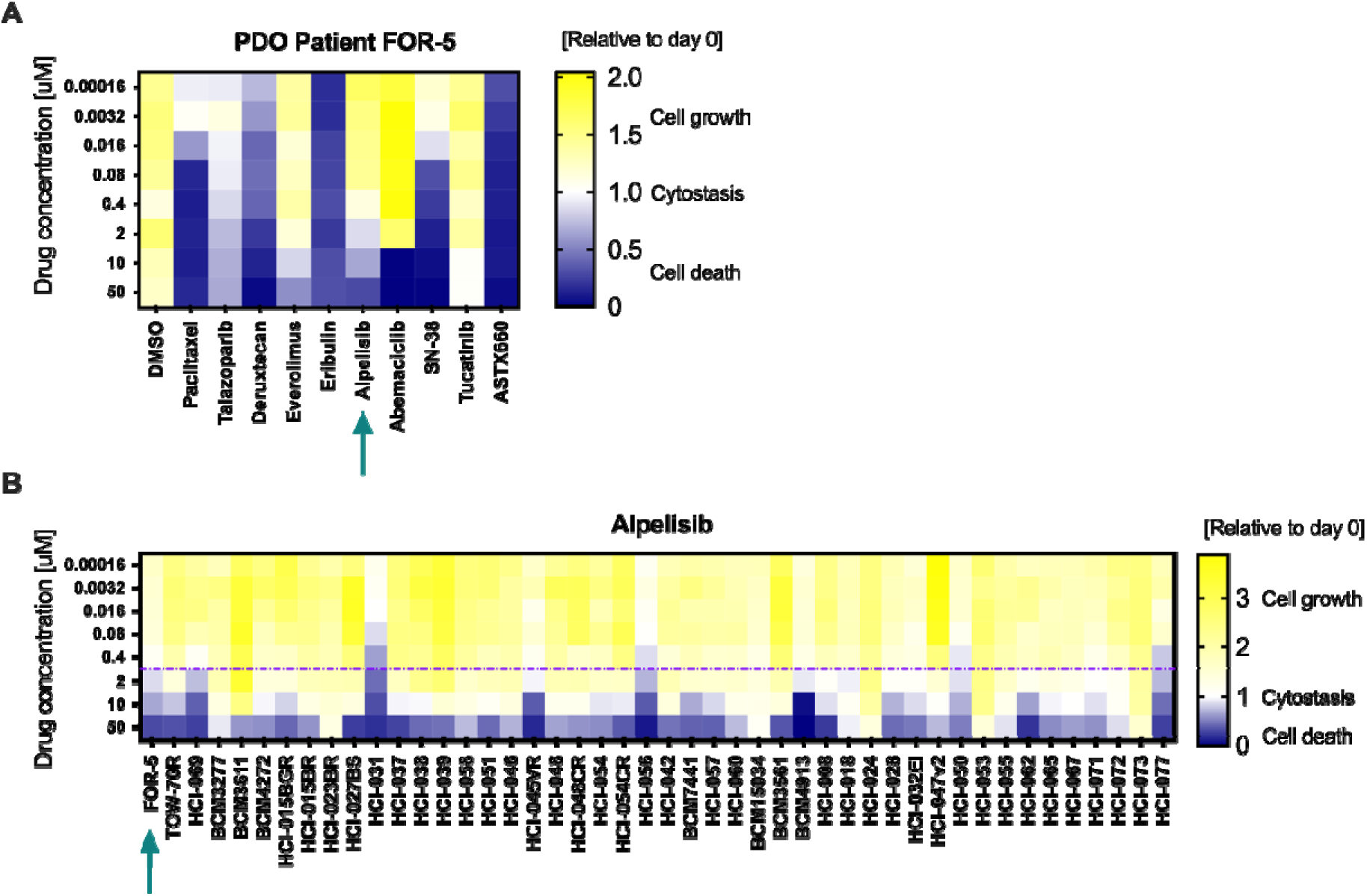
Pertinent FPO results that informed treatment selection for patient 5 (FOR-5). (A) PDO drug response assay results for patient FOR-5. The arrow indicates the nominated therapy, which also considered mutation status. (B) FOR-5 PDO drug sensitivity to alpelisib compared to responses in other patient-derived organoids previously tested. Dashed line indicates effective drug dose in FOR-5 PDO. Arrow indicates FOR-5 PDOs.

In the third PDO(+) case (Patient 6, Figure 6), on the basis of detection of a somatic *BRCA1* mutation, ctDNA nominated a PARP inhibitor; however, this nomination did not align with the FPO recommendation because talazoparib had mediocre antitumor activity on the respective PDOs relative to past patient-derived models tested. The FPO nomination was abemaciclib, supported by the high copy number gains in *CDK4* and *CDK6* on investigational tumor DNA testing. The prior diagnosis of weakly HR+/HER2- brain metastasis also factored into the decision to pursue abemaciclib (Tolaney et al., 2020). Even though the patient’s PFS2 was 3 months due to a weak antitumor response to abemaciclib and progression in the predominant systemic metastatic site (axillary lymph node), the brain metastases did not recur throughout her disease.

**Figure 6.**
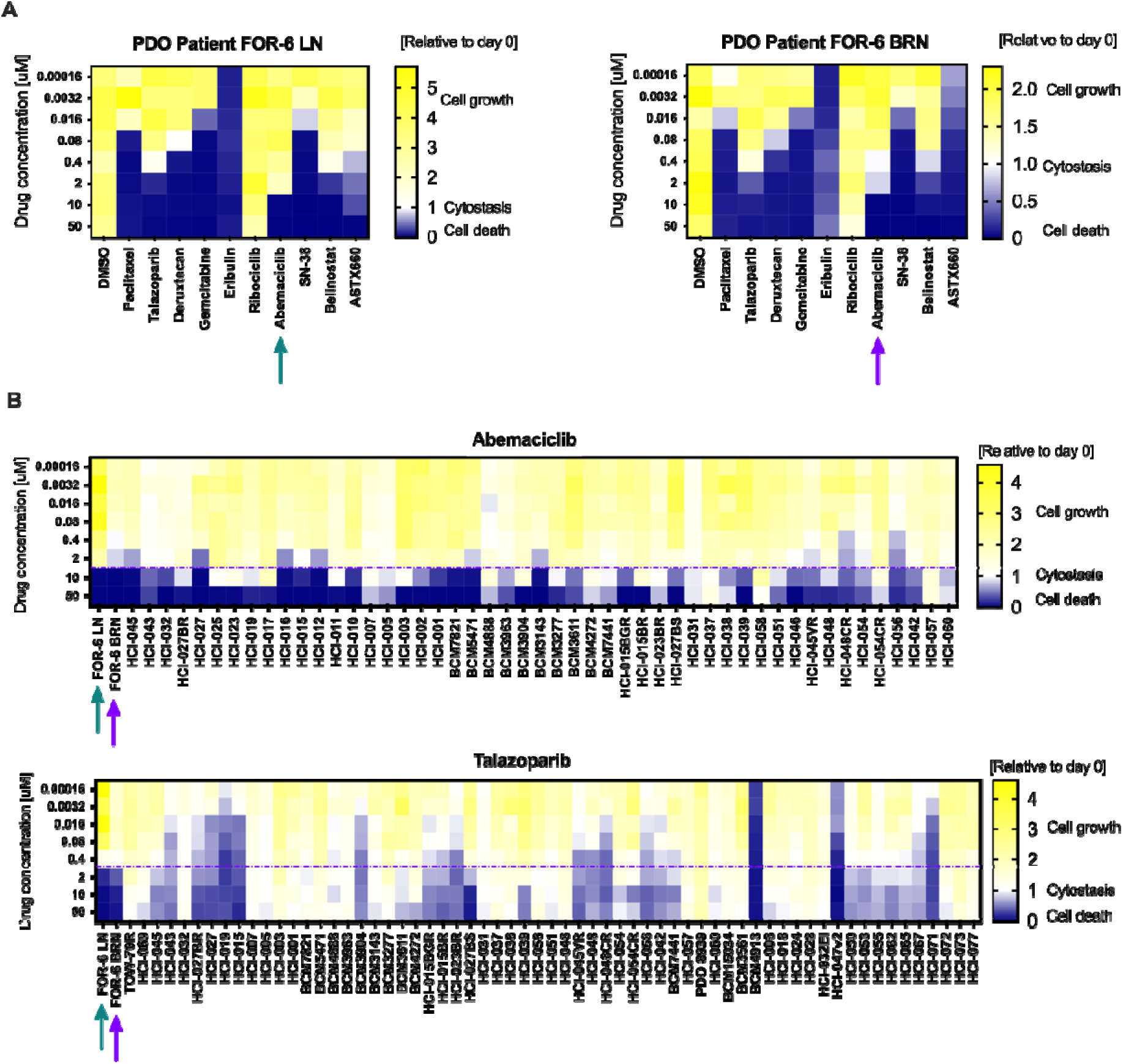
Pertinent FPO results that informed treatment selection for patient 6 (FOR-6). (A) drug response of patient FOR-6 PDOs, generated from an axillary lymph node biopsy (left panel; FOR-6 LN) and from a brain metastasis (right panel; FOR-6 BRN). PDO assays and mutation testing nominated abemaciclib (green arrow). Sensitivity of abemaciclib in FOR-6 BRN indicated by purple arrow. (B) Sensitivity of organoids of FOR-6 LN (green arrow) and FOR-6 BRN (purple arrow) to abemaciclib (upper panel, nominated by investigational genomics and FPO) and talazoparib (lower panel, nominated by ctDNA testing) compared to responses of other patient-derived organoids tested with abemaciclib and talazoparib, respectively. Dashed lines indicate cytotoxic drug dose in FOR-6 LN PDO relative to other models tested.

In the fourth PDO(+) case (Patient 12, Figure 7), based on detection of both *ESR1* and *PIK3CA* hotspot mutations on commercial ctDNA testing, elacestrant or fulvestrant in combination with alpelisib were nominated without prioritizing one option over the other. Investigational tumor DNA sequencing identified additional aberrations along the PI3K-AKT1 -mTOR pathway downstream of the mutated PIK3CA (amplification of AKT1 and AKT3, and loss of STK11, a known mTOR pathway suppressor), suggesting that a PI3K inhibitor would have not been effective. Note that elacestrant was not evaluated on the organoids. By the time that the *ESR1* mutation was detected, the patient had received 3 prior lines of endocrine combination therapy with a progression-free interval on the immediate prior line of therapy of only 2.3 months; exploratory analyses have shown that with such a short progression-free interval with the prior line of therapy, the benefit of elacestrant would have been small (Bardia et al., 2024). The FPO nomination was everolimus, the patient was rechallenged with exemestane and everolimus yielding a PFS2 of 1.75 months.

**Figure 7.**
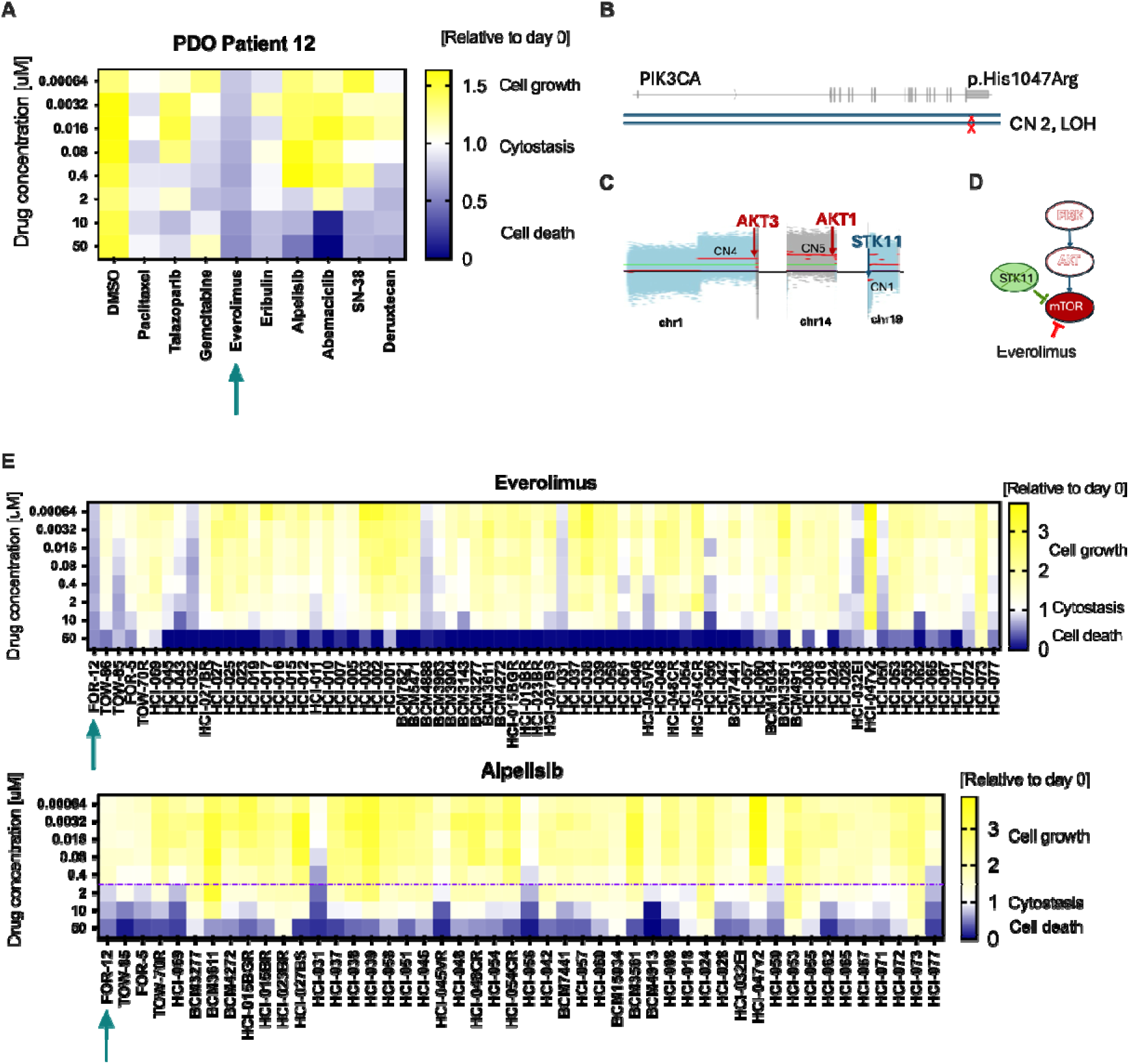
Pertinent FPO results that informed treatment selection for patient 12 (FOR-1 2). (A) PDO drug response assay results for FOR-12 PDOs with sensitivity for everolimus. Arrow indicates nominated therapy. (B) Genomic testing revealed PIK3CA mutation, (C) AKT1, AKT3 amplification and loss of STK11 (D) mTOR pathway regulation based on genomic testing results (E) FOR-12 PDO sensitivity to everolimus (upper panel, nominated by FPO and supported by investigational genomics) and alpelisib (lower panel, nominated by ctDNA testing) compared to responses in other patient-derived organoids tested. Arrows indicate FOR-12 PDOs. Dashed line indicates cytotoxic alpelisib dose in FOR-12 PDO relative to other models tested (top of graph for everolimus).

### Efficacy of the FPO-informed therapy

In all cases, PFS achieved with the informed therapy (PFS2) did not exceed the PFS achieved with the immediate prior therapy (PFS1) (Figure 3). PFS2/PFS1 ratios ranged between 0.26 – 0.6. It is worth noting that in 2 cases, the informed therapy constituted the final line of treatment that the patient received; in another case, the patient went on to receive one additional therapy following the informed therapy with a PFS of ∼4 months. The median overall survival (time from tum r tissue collection to death) for the PDO(+) patients was 10.1 months.

### Studies on longitudinally collected samples

We were able to obtain longitudinal tumor biopsies, generate the requisite PDOs, and evaluate their respective drug sensitivities from a PDO(+) patient with TNBC (Patient 6) who went on to receive 3 subsequent lines of therapy following her informed therapy (Figure 8, upper panel). In this patient, longitudinal PDOs showed similar (but not identical) responses to a limited five-drug panel. In 3 PDO(+) patients in whom commercial ctDNA testing was performed at the time of the investigational biopsy and repeated when the FPO-informed therapy was initiated, no new mutations were identified and the variant allele frequency of the already detected mutations increased (Figure 8, lower panel).

**Figure 8.**
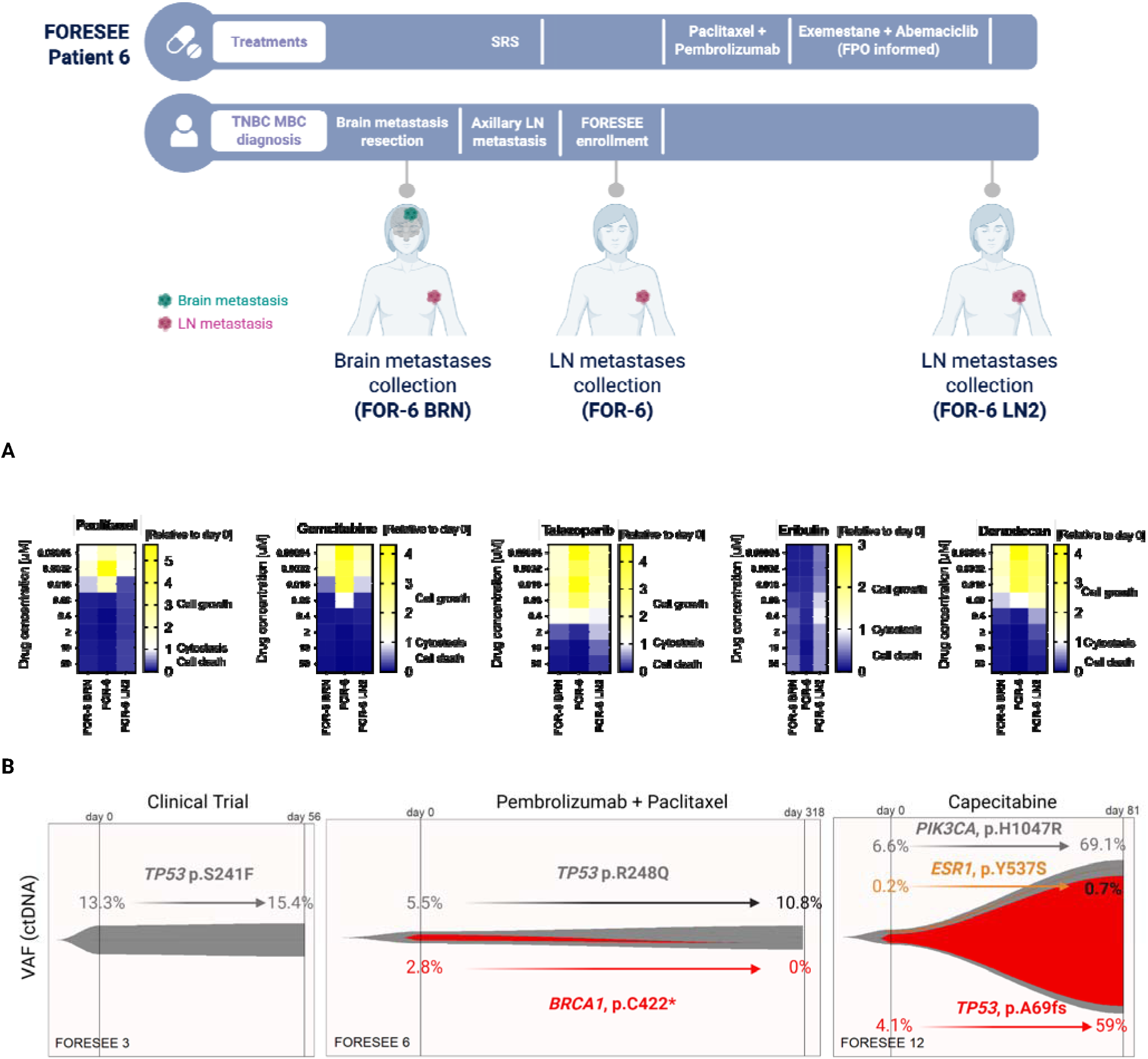
**Studies on longitudinally collected samples**. The natural history of metastatic disease for patient 6 (FOR-6) in whom longitudinal samples were obtained and PDOs developed. Tumor tissue from a brain metastasis (FOR-6 BRN), untreated lymph node metastasis (FOR-6) and previously treated with first line followed by the informed line of therapy lymph node metastasis (FOR-6 LN2). SRS, stereotactic radiosurgery to brain. (A) Dose response of longitudinally collected PDOs across 5 selected therapies. (B) Mutations detected on circulating tumor DNA and their respective variant allele frequency (VAF) at the time of registration/initiation of therapy preceding the FPO-informed therapy (day 0) vs. the time that the FPO-informed therapy was initiated (day 56, 318, 81, respectively) in 3 PDO(+) patients in whom commercial ctDNA was collected longitudinally. This corresponds to genomic changes occurring during PFS1. No new mutations were identified; the VAF of the already detected mutations increased. The *BRCA1*, p.C422 mutation in patient 6 was detected only in her isolated brain metastasis, not the lymph node metastases. Definitive treatment of the isolated brain metastasis with surgery and stereotactic radiation therapy led to the non-detection of this mutation upon retesting.

## Discussion

The current precision medicine paradigm, based on static genomic assessments, has limitations in successfully treating patients with metastatic breast cancer. Since studies utilizing patient-derived models hold promise to address these limitations, a *prospective* functional precision oncology trial, FORESEE, was conducted to systematically investigate whether our organoid-based FPO platform could be integrated into the clinical workflow and optimize treatment selection. The primary endpoint of the FORESEE trial was to determine if PDO-based functional drug screening, combined with comprehensive genomic characterization, would identify clinically actionable outcomes in more than 46% of patients. This endpoint was not met; we identified clinically actionable outcomes in only 40% of patients. While multiple studies, including our own, have reported that patient-derived models can emulate the disease of the originating patient (Hidalgo et al., 2014, Zhang et al., 2013), and observational trials have shown concordance of ex vivo drug testing with patient responses in the clinic (Wu et al., 2024; Zheng et al., 2025; Ooft et al., 2021; Jensen et al., 2023; Pamarthy and Sabaawy 2021), delivering the right drug to the right patient at the right time remains an elusive proposition - especially for very advanced disease.

The principal constraint that undermined the feasibility endpoint for FORESEE was PDO generation. Despite careful consideration of the target metastatic site for biopsy, low tumor content of the tissue collected sometimes led to low input material, compromising successful PDO establishment. Even with adequate input material, metastases from certain sites (particularly liver) as well as tumors with more indolent biology did not establish into stable PDOs; the biologic determinants of these failures are unknown. Interestingly, patients from whom PDOs were successfully established had a median overall survival of only 10.1 months. By comparison, patients from whom adequate metastatic tumor material was obtained from non-liver sites but PDO growth was not successful (n = 4), had a median overall survival of 25.9 months. Thus, growth of a tumor into PDO under current protocols may typify intrinsically aggressive treatment-refractory biology akin to tumor engraftment in xenografts (Vaklavas et al., 2024).

FORESEE was prospectively powered to test workflow feasibility rather than efficacy. The 1 00% utilization rate of FPO data, when it was available, by physicians in this trial should be interpreted as evidence that integrated FPO reports can be incorporated into clinical reasoning, not as evidence that they improve patient outcomes. A conceptual limitation of this feasibility study is that physician utilization and clinical benefit are not interchangeable endpoints.

The exploratory endpoint for clinical benefit was prespecified as PFS2/PFS1, which is a pragmatic, self-controlled metric suitable for small feasibility cohorts. None of the FPO-informed therapies in this trial met the prespecified exploratory endpoint of efficacy (PFS2/PFS1 > 1). However, early non-progression discontinuations and late-line initiation likely biased PFS2 downward. We did not perform censoring-based or time-to-treatment-discontinuation sensitivity analyses, so interpretation is limited by these late-line treatment and non-progression discontinuations. In at least two cases (Patients 3 and 5) the FPO nomination coincided with the standard-of-care option that would plausibly have been chosen without the report. Therefore, the degree to which the report altered rather than confirmed clinical decisions cannot be determined from this dataset. While it is unknown how other therapies would have fared, the FPO-informed therapies often constituted the optimal approved treatment option at that time. A limitation of our study was that it was not conducted in a CLIA-certified setting, so we could only return information to the physician for therapies that are FDA approved for breast cancer. The focus of this trial on simply prioritizing treatments that are already available to the patient may have also impacted negatively the efficacy endpoint of this study, because existing FDA-approved therapies are known to have limited efficacy in this difficult-to-treat population. PDOs established in the context of this trial will therefore constitute excellent models for future research to investigate novel therapeutic approaches.

To overcome the limitation of slow PDO generation for our FPO assays, we are currently conducting the TOWARDS-II clinical trial (NCT05464082) whereby patient-derived models are generated early and across the treatment course of HER2-breast cancer patients: at diagnosis of early-stage disease, at time of definitive surgery after preoperative chemotherapy (if there is residual disease), and at time of recurrence. This allows FPO results to be available prior to the diagnosis of recurrence to inform treatment selection. This strategy previously proved successful in a patient whose treatment selection upon recurrence of her triple negative metaplastic breast cancer was informed by organoids established at diagnosis of early stage disease (Vaklavas et al., 2024). Later, upon genomic profiling of the residual tumor after preoperative chemotherapy for early stage disease revealed a *PIK3CA* H1 047R mutation and organoid drug profiling predicted response to alpelisib. Upon metastatic recurrence, she enrolled in the FORESEE trial but a PDO from the pulmonary metastasis could not be established due to inadequate tumor content, so information from the prior sample was used. Despite prolonged interruptions in her treatment with alpelisib due to toxicity, her overall survival with alpelisib alone following the diagnosis of metastatic recurrence exceeded 14 months.

Alternative FPO strategies are currently being explored that allow for immediate drug testing obviating the need for organoid establishment such as *ex vivo* tissue slice cultures (Ladan et al., 2022). While *ex vivo* tissue slice cultures enable rapid drug screening, notable limitations include the requirement for large amounts of tumor material, and the application only to solid tumor samples; fluid-derived samples such as pleural effusions or ascites are not amenable to such testing. Quantitative phase imaging (QPI) is another functional strategy that offers rapid turnaround times for drug response assessment with single cell resolution and requirement for minimal input material (Polanco et al., 2026). An additional advantage of QPI is the ability to capture additional parameters of treatment response including depth and heterogeneity of response, features that can be particularly informative in dissecting tumor persistence and differential response in heterogeneous tumor populations (Polanco et al., 2022).

In summary, a FPO approach that entails PDO generation to inform treatment selection in metastatic HER2-breast cancer was limited by the low success rate of PDO establishment. However, when PDO were successful, drug responses were always obtained and were incorporated into the clinical decision rationale in all four cases, although the FPO-nominated therapy coincided with the standard-of-care or genomics-based option in a subset of cases, and no FPO-informed regimen achieved the exploratory efficacy endpoint of PFS2/PFS1 > 1. Tumor growth into PDO may constitute an intrinsic property of aggressive tumors for which currently available drugs are inadequate and novel therapeutic approaches are warranted. Methodological optimization of the patient-derived model generation, generation of PDOs earlier and across the natural history of breast cancer, and elucidation of the determinants of patient derived model generation can address the limitations of this trial.

## Methods

### Study design, eligibility and statistical considerations

The study was initiated after approval from the Institutional Review Board of the University of Utah (IRB #131027) and was conducted in accordance with the Declaration of Helsinki. Written informed consent was obtained from both patients and treating physicians given the investigational nature of the information shared with the physicians. Physicians were required to complete a pre- and post-information survey to capture how the drug screening and genomic results may affect the treatment selections made by the physicians by evaluating potential treatment plans prior to and after receiving results. If the patients went on to receive an investigational therapy, a separate consent for the respective clinical trial was required.

The primary endpoint was the proportion of cases where clinically actionable outcomes (defined as clinically meaningful therapeutic vulnerabilities to FDA approved medication(s) or investigational regimens being evaluated in ongoing separate clinical trials within Huntsman Cancer Institute, i.e. therapies accessible to the patient) identified by functional precision oncology by 12 weeks. Based on our preliminary data, we hypothesized that we would be able to establish PDO and return results in 46% of cases. A one-sided exact binomial test with null hypothesis that the true proportion of cases where clinically actionable outcomes identified by functional precision oncology by 12 weeks is 46% or lower was performed. The alternative hypothesis was that the proportion is higher than 46%. The primary endpoint proportion is reported here along with an exact binomial confidence interval calculated using the Clopper-Pearson method.

Secondary objectives included assessment of the time required to return genomic characterization and drug screening results to treating physicians and assessment of the performance and concordance of our integrated functional genomic assays (drug screen with or without genomic characterization) against commercially available assays. Assessment of the clinical efficacy of treatment decisions informed by functional genomic assays compared with treatment decisions not informed by functional genomic assays constituted an exploratory objective.

### Commercial genomic testing (ct/tissue)

The commercial partner for this trial was TEMPUS. The commercial tests performed in this clinical trial were the xT CDx (an FDA-approved 648-gene tissue-based matched tumor-normal NGS test for molecular profiling of all malignant solid tumors) and the xF (a 1 05-gene liquid biopsy NGS test that detects oncogenic drivers and resistance mutations, identifies single nucleotide variants, insertions and deletions, copy number gains, and gene rearrangements). xF also reports on blood tumor mutational burden and microsatellite instability-high status. Note that at the time that the trial was conducted, the xT test did not entail RNA sequencing.

### Tissue collection and PDO culture

In cases with solid tissue collection, 2-6 patient tumor biopsies were obtained, and cut into 3 mm long pieces. One biopsy piece was fixed in 1 0% NBF for pathohistological analysis, and one biopsy piece was flash frozen and stored at -80°C for genomic analysis. All remaining biopsy pieces were digested using gentle MACS Octoheater and the human tumor dissociation kit. Specifically, in a C-tube (cat# 1 30-096-334, Miltenyi Biotec), 5 ml breast PDO base media (Sachs et al., 2018) were mixed with human tumor dissociation kit enzymes (cat#, 130-095-929 Miltenyi Biotec) 100 ul R, 200 ul H, and 25 ul A enzymes and 5 nM Y-27632 (cat#, S1 049 Selleck Chemicals) was added. Tissue was digested in a gentle MACS Octoheater using the gentle MACS program 37C_h_TDK_3, and digested material was washed and embedded into a 200 ul matrigel dome in a 6 well plate as previously described (Scherer et al., 2023). To maximize tumor cell recovery the original biopsy tissue collection media was also saved, centrifuged and the pellet was either combined with the pellet of digested tissue fragments to embedded in the one dome or embedded separately in different dome, depending on the amount of original tissue used to generate the PDOs. Organoids were cultured with 5 ml of breast PDO complete medium (Sachs et al., 2018) with freshly added 5 nM Y-27632 and RSPO3 (cat# 120-44, PeproTech). In cases of thoracentesis specimens, the pleural effusion sample was centrifuged for 5 minutes (1800 rpm, 4 degrees). The cell pellet was resuspended, subjected to CD45 depletion (cat# 17898, Stemcell Technologies), a small fraction of the cells was reserved for genomic sequencing, and the remainder was embedded into matrigel domes as described above. For all PDO cultures, medium was exchanged every 3-4 days, and PDO cultures were expanded and passaged. Stroma and dead cells were removed via differential centrifugation (Scherer et al., 2023).

### PDO validation

To confirm tumor content in the PDO cultures, DNA was extracted from whole PDO domes or cell-free DNA (cfDNA) was isolated from conditioned media and subjected to copy-number variation (CNV) analysis (Supplemental Figure 1). 200-500 ng DNA was used for assays with Illumina Omni2.5Exome or Illumina Cyto+GDA chips by the Genomics Core Facility at the University of Utah. PDOs were further validated using short tandem repeat (STR) assay as quality control to ensure patient match. When sufficient material was available, PDO cultures were further characterized by RT-qPCR to assess epithelial content, hormone receptor expression, and the expression of mesenchymal stem cell and fibroblast markers (Supplemental Figure 2) as previously described (Guillen et al., 2022; Scherer et al., 2023) for the following gene products: *ESR1, PGR, ERBB2, TFF1, AR, GATA3, PDPN, CDH1, EPCAM, KRT8, TRPS1, SCGB2A2, CD246, CD1 40a, FAP, CD90, CD106, PTPRC, GAPDH*). Mesenchymal stem cell and fibroblast markers in validated PDO cultures showed absent or minimal expression, consistent with minimal stromal content in the PDO cultures at the time of drug screening.

### Nucleic acid extraction from solid tissue and conditioned media

Small pieces (∼1 mm^3^) from patient biopsy samples were either directly used for RNA/DNA extraction, or flash frozen and stored at -80°C. Tissues were lysed in RLT Buffer Plus with BME added freshly in a Qiagen Tissue Lyser II (3 x rounds: frequency 20/sec for 30 sec), followed by nucleic acids extraction with the Qiagen RNA/DNA Allprep micro kit (cat# 80284, Qiagen) according to the manufacturer’s instructions. Nucleic acids from PDOs were extracted using the Qiagen RNA/DNA Allprep mini kit (cat# 80204, Qiagen) in cases of whole dome extractions following published protocol (Scherer et al., 2023). When DNA was extracted from conditioned media (CM), a cell free DNA (cfDNA) extraction kit was used (cat# A2931 9, Thermo Fisher). Briefly, culture medium was conditioned for 4 days on PDO culture, and 5 ml of fresh or frozen CM was used for the extraction. In a 15ml conical tube, 75 ul Proteinase K (20 mg/ml, cat#03115828001, Sigma), 5 ml CM, and 250 ul SDS (20% solution, cat# 7990-OP, Millipore) was added in this order, mixed well and incubated in a water bath at 60°C for 20 min. Samples were cooled on ice for 5 min. For each sample, a master mix of 75 ul well-vortexed MagMAX Cell Free DNA Magnetic Beads was added to 6.25 ml MagMAX Cell Free DNA Lysis/Binding solution. The CM sample was vortexed, then magnetic bead solution was added, and thoroughly mixed by inverting tube 10 times. The tube was vigorously shaken on the vortex for 25 min at RT. The sample was placed on DynaMAG1 5 magnet (Thermo Fisher #1 2301 D) for 5 min until solution cleared. The supernatant was carefully aspirated, the tube was removed from the magnet and the magnetic beads were resuspended in 1 ml of MagMAX Cell Free DNA Wash Solution, and transferred to a 1.7 ml microcentrifuge tube. Sample was vortexed for 30 sec, then placed on a DynaMAG 2 magnet (Thermo Fisher #1 2321 D) for 2 min until solution cleared. The supernatant was removed, and the washing step was repeated for a total of two washes with 1 ml of MagMAX Cell Free DNA Wash Solution, and two washes with 1 ml of 80% ethanol (DNase/RNase-free). After the last wash, all supernatant was removed and the beads were left to dry at RT for 5 min until all liquid had evaporated. The sample was eluted by adding 60 μL Elution Buffer, vortexing for 5 min at max speed, and placing tube on the magnet for 2 min. The DNA-containing supernatant was transferred into new tube and quantified using Qubit HS dsDNA kit (cat# Q32851, Thermo Fisher Scientific).

### RNA/DNA sequencing and single cell/nuclei sequencing

Libraries for bulk RNA sequencing and whole genome sequencing (WGS) were prepared and sequenced at the Huntsman Cancer Institute High-Throughput Genomics and Bioinformatics Core. In cases of WGS, germline DNA obtained from the patient’s blood collection was submitted together with the patient’s tumor DNA. For single cell or single nuclei sequencing, cryopreserved samples were thawed at 37 °C for 5 min and immediately transferred to low-binding microcentrifuge tubes. Cells were pelleted by centrifugation, the supernatant was aspirated, and the pellet was gently resuspended in 1 000 μL of Accutase. Samples were incubated for 15 min at room temperature to promote dissociation. Following enzymatic treatment, cells were centrifuged and washed with 250 μL of PBS containing 1 % BSA. A second wash was performed under the same conditions, after which cells were counted and viability was assessed. Dead-cell removal was performed if viability fell below 50%. For nuclei preparation, cytoprotective reagents were prepared fresh on the day of use and maintained at 4 °C. Tissue samples were thawed in 0.5 ml of ice-cold phosphate-buffered saline (PBS; devoid of Ca²⁺ and Mg²⁺), to which the following supplements were added: 0.54 μM Necrostatin (Inc. #J65341, ThermoFisher Scientific Chemicals, Waltham, MA), 1 μM HPN-07 (SML21 63, Sigma-Aldrich), 0.32 μM sodium hydroxybutyrate (A11 61 31 4, ThermoFisher Scientific Chemicals), 78 nM Q-VD-Oph (SML0063, Sigma-Aldrich, St. Louis, MO), and 0.2 U/ml of Ribolock RNase inhibitor (EO0381, Life Technologies Corporation, Carlsbad, CA). Tissue was transferred to a 2-mL Dounce homogenizer containing 1.5 mL of nuclei extraction buffer supplemented with 0.2 U/ml RNase inhibitor, and homogenized using 10 strokes with pestle A followed by 10 strokes with pestle B. The suspension was incubated on ice for 5 min, subjected to an additional set of pestle B strokes, incubated again for 5 min, and homogenized once more with pestle B. Nuclear suspensions were centrifuged at 600 × g for 5 min in a swinging-bucket rotor, and pellets were gently resuspended in PBS with 2% BSA and RNase inhibitor. Final suspensions were sequentially filtered through 70-μm and 40-μm FLOWMI strainers to obtain a clean, uniform nuclear preparation for single-nuclei sequencing.

### Sequencing data analysis

#### WGS Data Processing and Somatic Variant Calling

Sequencing reads in their raw form were mapped to the GRCh38 human reference genome using BWA-MEM (v0.7.17). Detection of single-nucleotide variants (SNVs) and short insertions/deletions (indels) was carried out with FreeBayes (v1 .3.4). To retain only high-confidence variants, the following filtering criteria were applied: 1) a minimum variant quality score of 20; 2) a minimum per-sample sequencing depth of 15 and a minimum per-tumor-sample depth of 30; 3) an allele frequency (VAF) exceeding 0.05 across all samples; 4) overlap with the 1000 Genomes Project genome accessibility mask for GRCh38; and 5) exclusion of multi-allelic sites. Variants were classified as somatic when no variant-supporting reads were present in the matched normal control sample. Functional annotation of SNVs and short indels was performed using SnpEff (v4.3). Somatic copy number alterations (CNAs) and loss of heterozygosity (LOH) were determined using FACETS (v0.6.1). This tool leverages the tumor-to-normal depth ratio to infer copy number changes in the tumor, and utilizes the ratio of alternative to reference allele counts (odds ratio) at single nucleotide polymorphism sites within the tumor to detect LOH and allelic imbalance.

#### Single Cell RNAseq Data Processing and Analysis

Single-cell libraries were constructed on the 1 0x Genomics Chromium platform using the 1 0X 3’ v3.1 chemistry with unique dual indices, following the manufacturer’s guidelines, with a target capture of 6,000 cells per sample. Sequencing was performed on an Illumina platform, aiming for at least 400 million read pairs per library. The raw sequencing output was processed through Cell Ranger (v7), which handled alignment to the GRCh38 human reference genome and generation of gene expression count matrices. Count matrices from multiple samples were combined using the Cell Ranger “aggr” command without applying read depth normalization.

#### Single-Cell Data Analysis and Tumor Cell Identification

The filtered count matrices were analyzed with Seurat. A quality control step was carried out to exclude low-quality cells using the following thresholds: cells expressing fewer than 200 genes and cells in which mitochondrial reads exceeded 25% of total reads. Normalization was performed via scTransform, and highly variable genes were selected for dimensionality reduction. Principal component analysis (PCA) was conducted, and the top 30 components were used as input for both uniform manifold approximation and projection (UMAP) visualization and graph-based clustering via the nearest neighbor algorithm. Cell types were annotated based on established marker genes: immune cells were identified by PTPRC; stromal cells by ALDH1 A2; fibroblasts by COL3A1, FBN1, LUM, and VIM; and tumor cells by EPCAM.

#### Vulnerability Annotation

Genes harboring mutations (identified through WGS) and genes showing differential expression (identified through scRNA-seq) were cross-referenced with the COSMIC Cancer Gene Census (Sinkala et al., 2023) and the TARGET database of actionable genomic vulnerabilities (Van Allen et al., 2014). Pathogenic mutations, high-level copy number gains (greater than 4 copies), and deletions were mapped at the gene level. The TARGET database was consulted to determine whether these genomic alterations are associated with known indicators of tumor sensitivity or resistance to specific drug classes.

### PDO drug testing

PDO cultures with confirmed tumor content were objected to a 6-day drug testing using the published method (Guillen et al., 2022; Scherer et al., 2023) with CTG3D as readout with the exception that the assay was performed using breast PDO complete media (Sachs et al., 2018) to keep media consistent throughout the PDO culture and drug testing. The drug panel for each patient was carefully curated together with the treating clinician to ensure any tested drug would be available to the patient.

## Data Availability

All data produced in the present study are available upon reasonable request to the authors.

## Supplemental Figures and Tables

**Supplemental Figure 1.**
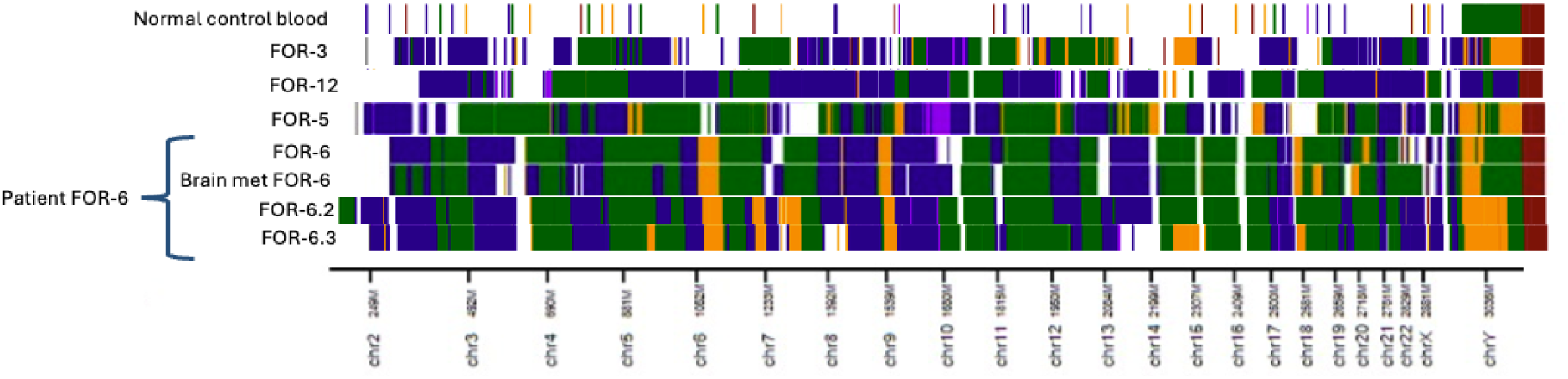
CNV profiling of PDOs confirms tumor content.

**Supplemental Figure 2.**
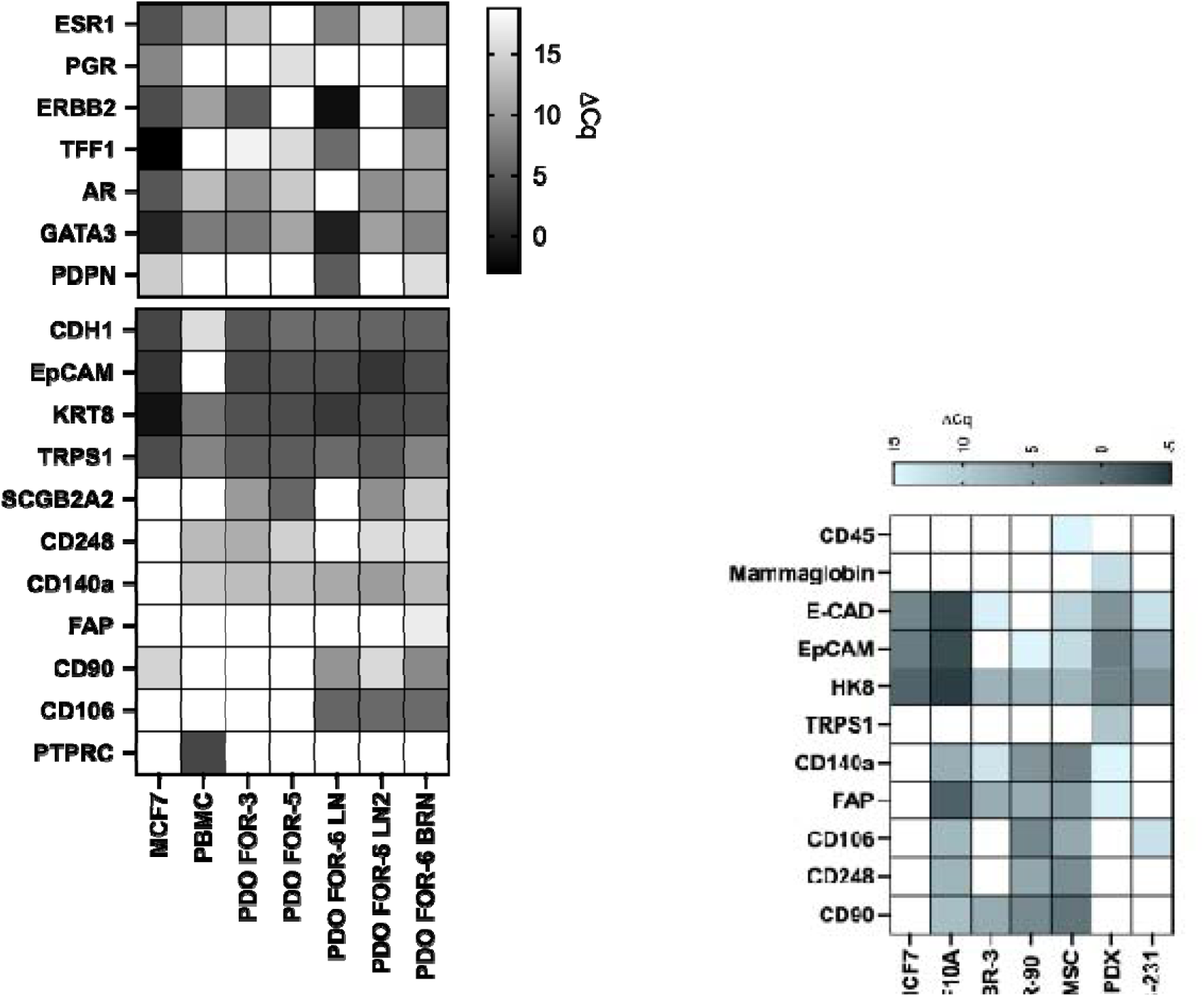
**qRT-PCR profiling of PDOs**. Gene expression of PDOs (left), and cell lines and PDX lines used as controls for gene expression panel (right).

**Supplemental Table 1.**

|  |  |
| --- | --- |
|  | Enrolled patients (n=15) |

|  | Enrolled patients (n=15) |
| --- | --- |
| **HR+/HER2-** | 8 |
| Age (median, range) | 52 (41.7 - 61) |
| Relapsed (n) | 5 |
| De novo (n) | 3 |
| Number of prior lines of therapy (median, range) | 3.5 (1 - 5) |
| Metastatic sites (median, range) | 3 (1 - 6) |
| bone | 8 |
| liver | 6 |
| Median time on therapy immediately prior to enrollment (months) | 2.27 |
| Germline pathogenic mutation (n=7)* | 2 |
| **TNBC** | 7 |
| Age (median, range) | 57 (37.3 - 86.3) |
| Early relapse (<1 year, n) | 2 |
| Late relapse (>1 year, n) | 4 |
| De novo (n) | 1 |
| Metastatic sites (median, range) | 2 (1 - 3) |
| Visceral involvement | 4 |
| Germline pathogenic mutation** | 3 |

\*Germline pathogenic variants in *BRCA2* (n=1) and *MUTYH* (n=1)
\*\*Germline pathogenic variants in *BRCA2* (pathogenic/likely pathogenic, n=1), *MUTYH* (n=1), *ATM* (n=1).

## Acknowledgements

We gratefully acknowledge the patients who participated in this study, as well as expertise from the coordinators in the Clinical Trials Office at Huntsman Cancer Institute. Research reported in this publication utilized the High-Throughput Genomics Resources at Huntsman Cancer Institute at the University of Utah, and the Genomics Core Facility at the University of Utah. Images were created using BioRender.com.

## Author Contribution Statement

C.V., S.S.B., and K.K. ran the trial. S.D.S, L.Z., C-H. Y., E. C-S., A.B., X.H., Y.Q., and P.J.M. performed experiments and data analysis. K.M.B. performed statistical analysis. C.V., G.M., B.E.W., and A.L.W. supervised the work and secured funding. C.V, S.D.S, and A.L.W wrote the manuscript. All authors reviewed and edited the manuscript.

## References

Gladstone BP, Beha J, Hakariya A, Missios P, Malek NP, Bitzer M. Systematic review and meta-analysis of molecular tumor board data on clinical effectiveness and evaluation gaps. NPJ Precis Oncol. 2025 Apr 2;9(1):96. doi: 10.1038/s41698-025-00865-1. PMID: 40175535; PMCID: PMC11965449.

Schwartzberg L, Kim ES, Liu D, Schrag D. Precision Oncology: Who, How, What, When, and When Not? Am Soc Clin Oncol Educ Book. 2017;37:160–169. PMID: 28561651

Flaherty KT, Gray RJ, Chen AP, Li S, McShane LM, Patton D, Hamilton SR, Williams PM, Iafrate AJ, Sklar J, Mitchell EP, Harris LN, Takebe N, Sims DJ, Coffey B, Fu T, Routbort M, Zwiebel JA, Rubinstein LV, Little RF, Arteaga CL, Comis R, Abrams JS, O’Dwyer PJ, Conley BA, NCI-MATCH team. Molecular Landscape and Actionable Alterations in a Genomically Guided Cancer Clinical Trial: National Cancer Institute Molecular Analysis for Therapy Choice (NCI-MATCH). J Clin Oncol. 2020 Nov 20;38(33):3883–3894. PMID: 33048619

O’Dwyer PJ, Gray RJ, Flaherty KT, Chen AP, Li S, Wang V, McShane LM, Patton DR, Tricoli JV, Williams PM, Iafrate AJ, Sklar J, Mitchell EP, Takebe N, Sims DJ, Coffey B, Fu T, Routbort M, Rubinstein LV, Little RF, Arteaga CL, Marinucci D, Hamilton SR, Conley BA, Harris LN, Doroshow JH. The NCI-MATCH trial: lessons for precision oncology. Nat Med. 2023 Jun;29(6):1349–1357. doi: 10.1038/s41591-023-02379-4. Epub 2023 Jun 15. PMID: 37322121; PMCID: PMC10612141.

Hidalgo M, Amant F, Biankin AV, Budinská E, Byrne AT, Caldas C, Clarke RB, de Jong S, Jonkers J, Mælandsmo GM, Roman-Roman S, Seoane J, Trusolino L, Villanueva A. Patient-derived xenograft models: an emerging platform for translational cancer research. Cancer Discov. 2014 Sep;4(9):998–1013. doi: 10.1158/2159-8290.CD-14-0001. Epub 2014 Jul 15. PMID: 25185190; PMCID: PMC4167608.

Zhang X, Claerhout S, Prat A, Dobrolecki LE, Petrovic I, Lai Q, Landis MD, Wiechmann L, Schiff R, Giuliano M, Wong H, Fuqua SW, Contreras A, Gutierrez C, Huang J, Mao S, Pavlick AC, Froehlich AM, Wu MF, Tsimelzon A, Hilsenbeck SG, Chen ES, Zuloaga P, Shaw CA, Rimawi MF, Perou CM, Mills GB, Chang JC, Lewis MT. A renewable tissue resource of phenotypically stable, biologically and ethnically diverse, patient-derived human breast cancer xenograft models. Cancer Res. 2013 Aug 1;73(15):4885–97. doi: 10.1158/0008-5472.CAN-12-4081. Epub 2013 Jun 4. PMID: 23737486; PMCID: PMC3732575.

NCI-MATCH sets “benchmark of actionability.” Cancer Discov. American Association for Cancer Research (AACR); 2021 Jan;11(1):6–7. PMID: 33168587

Sachs N, de Ligt J, Kopper O, Gogola E, Bounova G, Weeber F, Balgobind AV, Wind K, Gracanin A, Begthel H, Korving J, van Boxtel R, Duarte AA, Lelieveld D, van Hoeck A, Ernst RF, Blokzijl F, Nijman IJ, Hoogstraat M, van de Ven M, Egan DA, Zinzalla V, Moll J, Boj SF, Voest EE, Wessels L, van Diest PJ, Rottenberg S, Vries RGJ, Cuppen E, Clevers H. A Living Biobank of Breast Cancer Organoids Captures Disease Heterogeneity. Cell. 2018 Jan 11;172(1-2):373–386.e10. doi: 10.1016/j.cell.2017.11.010. Epub 2017 Dec 7. PMID: 29224780.

Pauli C, Hopkins BD, Prandi D, Shaw R, Fedrizzi T, Sboner A, Sailer V, Augello M, Puca L, Rosati R, McNary TJ, Churakova Y, Cheung C, Triscott J, Pisapia D, Rao R, Mosquera JM, Robinson B, Faltas BM, Emerling BE, Gadi VK, Bernard B, Elemento O, Beltran H, Demichelis F, Kemp CJ, Grandori C, Cantley LC, Rubin MA. Personalized In Vitro and In Vivo Cancer Models to Guide Precision Medicine. Cancer Discov. 2017 May;7(5):462–477. doi: 10.1158/2159-8290.CD-16-1154. Epub 2017 Mar 22. PMID: 28331002; PMCID: PMC5413423.

Letai A, Bhola P, Welm AL. Functional precision oncology: Testing tumors with drugs to identify vulnerabilities and novel combinations. Cancer Cell. 2022 Jan 10;40(1):26–35. doi: 10.1016/j.ccell.2021.12.004. Epub 2021 Dec 23. PMID: 34951956; PMCID: PMC8752507.

Guillen KP, et al. A human breast cancer-derived xenograft and organoid platform for drug discovery and precision oncology. Nat Cancer 2022 Feb;3(2):232–250.

Chen P, Zhang X, Ding R, Yang L, Lyu X, Zeng J, Lei JH, Wang L, Bi J, Shao N, Shu D, Wu B, Wu J, Yang Z, Wang H, Wang B, Xiong K, Lu Y, Fu S, Choi TK, Lon NW, Zhang A, Tang D, Quan Y, Meng Y, Miao K, Sun H, Zhao M, Bao J, Zhang L, Xu X, Shi Y, Lin Y, Deng C. Patient-Derived Organoids Can Guide Personalized-Therapies for Patients with Advanced Breast Cancer. Adv Sci (Weinh). 2021 Nov;8(22):e2101176. doi: 10.1002/advs.202101176. Epub 2021 Oct 4. PMID: 34605222; PMCID: PMC8596108.

Scherer SD, et al. Breast cancer PDxO cultures for drug discovery and functional precision oncology. STAR Protoc 2023 Sep 15;4(3):102402.

Andre F, et al. Comparative genomic hybridisation array and DNA sequencing to direct treatment of metastatic breast cancer: a multicentre, prospective trial (SAFIR01/UNICANCER). Lancet Oncol. 2014 Mar;15(3):267–74.

Verstegen MMA, Coppes RP, Beghin A, De Coppi P, Gerli MFM, de Graeff N, Pan Q, Saito Y, Shi S, Zadpoor AA, van der Laan LJW. Clinical applications of human organoids. Nat Med. 2025 Feb;31(2):409–421. doi: 10.1038/s41591-024-03489-3. Epub 2025 Feb 3. PMID: 39901045.

Zheng M, Yao L, Jing N, Wang Y, Wang X, Yang J. Personalized Treatment for Invasive Ductal Breast Carcinoma with Lung and Liver Metastases Based on Patient-Derived Organoids: A Case Report. Onco Targets Ther. 2025 Oct 25;18:1189–1194. doi: 10.2147/OTT.S540143. PMID: 41169604; PMCID: PMC12570986.

Tolaney SM, Sahebjam S, Le Rhun E, Bachelot T, Kabos P, Awada A, Yardley D, Chan A, Conte P, Diéras V, Lin NU, Bear M, Chapman SC, Yang Z, Chen Y, Anders CK. A Phase II Study of Abemaciclib in Patients with Brain Metastases Secondary to Hormone Receptor-Positive Breast Cancer. Clin Cancer Res. 2020 Oct 15;26(20):5310–5319. doi: 10.1158/1078-0432.CCR-20-1764. Epub 2020 Jul 21. Erratum in: Clin Cancer Res. 2021 Mar 1;27(5):1582. doi: 10.1158/1078-0432.CCR-21-0193. PMID: 32694159.

Wu H, Wang W, Zhang Y, Chen Y, Shan C, Li J, Jia Y, Li C, Du C, Cai Y, Zhang Y, Zhang S, Wu F. Establishment of patient-derived organoids for guiding personalized therapies in breast cancer patients. Int J Cancer. 2024 Jul 15;155(2):324–338. doi: 10.1002/ijc.34931. Epub 2024 Mar 27. PMID: 38533706.

Ooft SN, Weeber F, Schipper L, Dijkstra KK, McLean CM, Kaing S, van de Haar J, Prevoo W, van Werkhoven E, Snaebjornsson P, Hoes LR, Chalabi M, van der Velden D, van Leerdam M, Boot H, Grootscholten C, Huitema ADR, Bloemendal HJ, Cuppen E, Voest EE. Prospective experimental treatment of colorectal cancer patients based on organoid drug responses. ESMO Open. 2021 Jun;6(3):100103. doi: 10.1016/j.esmoop.2021.100103. Epub 2021 Apr 19. PMID: 33887686; PMCID: PMC8086019.

Jensen LH, Rogatto SR, Lindebjerg J, Havelund B, Abildgaard C, do Canto LM, Vagn-Hansen C, Dam C, Rafaelsen S, Hansen TF. Precision medicine applied to metastatic colorectal cancer using tumor-derived organoids and in-vitro sensitivity testing: a phase 2, single-center, open-label, and non-comparative study. J Exp Clin Cancer Res. 2023 May 5;42(1):115. doi: 10.1186/s13046-023-02683-4. PMID: 37143108; PMCID: PMC10161587.

Pamarthy S, Sabaawy HE. Patient derived organoids in prostate cancer: improving therapeutic efficacy in precision medicine. Mol Cancer. 2021 Sep 29;20(1):125. doi: 10.1186/s12943-021-01426-3. PMID: 34587953; PMCID: PMC8480086.

Cortes J, Rugo HS, Cescon DW, Im SA, Yusof MM, Gallardo C, Lipatov O, Barrios CH, Perez-Garcia J, Iwata H, Masuda N, Torregroza Otero M, Gokmen E, Loi S, Guo Z, Zhou X, Karantza V, Pan W, Schmid P; KEYNOTE-355 Investigators. Pembrolizumab plus Chemotherapy in Advanced Triple-Negative Breast Cancer. N Engl J Med. 2022 Jul 21;387(3):217–226. doi: 10.1056/NEJMoa2202809. PMID: 35857659.

Bardia A, Hu X, Dent R, Yonemori K, Barrios CH, O’Shaughnessy JA, Wildiers H, Pierga JY, Zhang Q, Saura C, Biganzoli L, Sohn J, Im SA, Lévy C, Jacot W, Begbie N, Ke J, Patel G, Curigliano G; DESTINY-Breast06 Trial Investigators. Trastuzumab Deruxtecan after Endocrine Therapy in Metastatic Breast Cancer. N Engl J Med. 2024 Dec 5;391(22):2110–2122. doi: 10.1056/NEJMoa2407086. Epub 2024 Sep 15. PMID: 39282896.

Bardia A, Cortés J, Bidard FC, Neven P, Garcia-Sáenz J, Aftimos P, O’Shaughnessy J, Lu J, Tonini G, Scartoni S, Paoli A, Binaschi M, Wasserman T, Kaklamani V. Elacestrant in ER+, HER2- Metastatic Breast Cancer with ESR1-Mutated Tumors: Subgroup Analyses from the Phase III EMERALD Trial by Prior Duration of Endocrine Therapy plus CDK4/6 Inhibitor and in Clinical Subgroups. Clin Cancer Res. 2024 Oct 1;30(19):4299–4309. doi: 10.1158/1078-0432.CCR-24-1073. PMID: 39087959; PMCID: PMC11443208.

Polanco, E.R., Moustafa, T.E., Balcioglu, O. et al. Determination of drug sensitivity in patient derived models of breast cancer by multiparametric quantitative phase imaging. BMC Cancer (2026). 10.1186/s12885-026-16752-2

Polanco ER, Moustafa TE, Butterfield A, Scherer SD, Cortes-Sanchez E, Bodily T, Spike BT, Welm BE, Bernard PS, Zangle TA. Multiparametric quantitative phase imaging for real-time, single cell, drug screening in breast cancer. Commun Biol. 2022 Aug 8;5(1):794. doi: 10.1038/s42003-022-03759-1. PMID: 35941353; PMCID: PMC9360018.

Liu Y, Liang S, Li J. Partial Response Achieved in a Case of Metastatic Triple-Negative Breast Cancer Based on the Patient-Derived Tumor Organoids. Int J Womens Health. 2025 Jul 31;17:2395–2400. doi: 10.2147/IJWH.S520867. PMID: 40765576; PMCID: PMC12323790.

Ladan MM, Meijer TG, Verkaik NS, Komar ZM, van Deurzen CHM, den Bakker MA, Kanaar R, van Gent DC, Jager A. Functional Ex Vivo Tissue-Based Chemotherapy Sensitivity Testing for Breast Cancer. Cancers (Basel). 2022 Feb 28;14(5):1252. doi: 10.3390/cancers14051252. PMID: 35267560; PMCID: PMC8909506.

Divoux J, Florent R, Jacobs M, Lequesne J, Grellard JM, San C, Grossi S, Kerdja K, Clarisse B, Boudier G, Cherifi F, Briand M, Dolivet E, Johnson A, Dubois B, Harter V, Lacroix J, Raboutet C, Marie B, Rousseau N, Blanc-Fournier C, Vaur D, Figeac M, Poulain L, Weiswald LB, Emile G. The TRIPLEX study: use of patient-derived tumor organoids as an innovative tool for precision medicine in triple-negative breast cancer. BMC Cancer. 2023 Sep 19;23(1):883. doi: 10.1186/s12885-023-11362-8. PMID: 37726786; PMCID: PMC10508010.

Vaklavas C, EcheMatsen CB, Chu Z, Boucher KM, Scherer SD, Pathi S, Beck A, Brownson KE, Buys SS, Chittoria N, D’Astous E, Gulbahce HE, Henry NL, Kimani S, Porretta J, Rosenthal R, Ward J, Wei M, Welm BE, Welm AL. TOWARDS Study: Patient-Derived Xenograft Engraftment Predicts Poor Survival in Patients With Newly Diagnosed Triple-Negative Breast Cancer. JCO Precis Oncol. 2024 Jul;8:e2300724. doi: 10.1200/PO.23.00724. PMID: 39074345; PMCID: PMC11371112.

Sinkala M. Mutational landscape of cancer-driver genes across human cancers. Sci Rep. 2023 Aug 7;13(1):12742. doi: 10.1038/s41598-023-39608-2. PMID: 37550388; PMCID: PMC10406856.

Van Allen EM, Wagle N, Stojanov P, Perrin DL, Cibulskis K, Marlow S, Jane-Valbuena J, Friedrich DC, Kryukov G, Carter SL, McKenna A, Sivachenko A, Rosenberg M, Kiezun A, Voet D, Lawrence M, Lichtenstein LT, Gentry JG, Huang FW, Fostel J, Farlow D, Barbie D, Gandhi L, Lander ES, Gray SW, Joffe S, Janne P, Garber J, MacConaill L, Lindeman N, Rollins B, Kantoff P, Fisher SA, Gabriel S, Getz G, Garraway LA. Whole-exome sequencing and clinical interpretation of formalin-fixed, paraffin-embedded tumor samples to guide precision cancer medicine. Nat Med. 2014 Jun;20(6):682–8. doi: 10.1038/nm.3559. Epub 2014 May 18. PMID: 24836576; PMCID: PMC4048335.

